# Cross-phenotype associations between Alzheimer’s Disease and its comorbidities may provide clues to progression

**DOI:** 10.1101/2023.11.06.23297993

**Authors:** Anni Moore, Marylyn D. Ritchie

## Abstract

Alzheimer’s disease (AD) is the most prevalent neurodegenerative disease worldwide, with one in nine people over the age of 65 living with the disease in 2023. In this study, we used a phenome wide association study (PheWAS) approach to identify cross-phenotype associations between previously identified genetic AD and for electronic health record (EHR) diagnoses from the UK Biobank (UKBB) (n=361,194 of European ancestry) and the eMERGE Network (n=105,108 of diverse ancestry). Based on 497 previously identified AD-associated variants from the Alzheimer’s Disease Variant Portal (ADVP), we found significant associations primarily in immune and cardiac related diseases in our PheWAS. Replicating variants have widespread impacts on immune genes in diverse tissue types. This study demonstrates the potential of using the PheWAS strategy to improve our understanding of AD progression as well as identify potential drug repurposing opportunities for new treatment and disease prevention strategies.

## Introduction

### AD remains prevalent in the aging population without effective treatment

Late onset Alzheimer’s disease (AD) remains one of the most rampant diseases in the aging population, with as many as 11% of people over the age of 65 living with AD^1^. AD is a neurodegenerative disease that is characterized by the deposition of amyloid-beta (Aβ) protein plaques around neurons and the appearance of tau tangles in the brain^2–4^. This ultimately leads to the death of these neurons which results in cognitive and motor deficits affecting patients’ everyday life. Biological changes that lead to AD have a substantial genetic component (h^2^=58-79%)^5^ and occur as many as 10-20 years before outward visible signs and symptoms^6^. Yet despite decades of research, no single treatment has been discovered or produced to cure AD, making identifying methods of early intervention and prevention even more important^7^.

### AD is a complex disease brought on by a combination of changes

Though single, high effect genetic variants account for approximately 5% of cases, more than 50% of AD cases are brought on by a combination of low-effect common variants^8^. AD has been linked to perturbations in many different biological pathways, often occurring simultaneously over the course of disease progression. Genetic variants associated with AD have been known to affect multiple biological systems including immune dysfunction, altered lipid metabolism, and neuroinflammation^9–11^. Additionally, a spectrum of other common, complex diseases have been associated with increased AD risk. For example, genetic variants impacting mitochondrial dysfunction and insulin signaling have been implicated in both type 2 diabetes (T2D) and AD^12,13,14^. In addition to T2D, cardiovascular disease, gastrointestinal (GI) dysfunction, and depression have all been associated with AD, often occurring well before an AD diagnosis^15–21^. Several known AD risk variants have been identified as pleiotropic between AD and other disorders^8,22,23^. All of these comorbidities suggest that broadly exploring pleiotropy between AD-associated variants/genes and other clinical phenotypes may lead to potential avenues for future therapeutic opportunities.

### Other diseases may provide clues to AD progression and therapeutics

With many of the aforementioned chronic diseases potentially affecting overall clinical status and progression of AD, they may also be used as notable risk factors in patients not yet diagnosed with AD. Identifying these shared biological pathways between AD and comorbid diseases may increase our understanding of AD pathology. Additionally, using genetics to identify causal variants that affect these overlapping pathways could provide a starting point to an alternative prevention strategy for those with preclinical-AD. This becomes especially relevant for comorbid diseases with existing treatments that target similar pathways that link them to AD progression. For example, insulin, which is used to treat patients with diabetes, has also been investigated for treatment for Alzheimer’s dementia. Pilot clinical trials have shown improvement in preserving general cognition in patients with cognitive impairment and AD ^24,25,26^. Further identification of diseases with overlapping pathways could be useful for drug repurposing. Here we use large medical biobanks to evaluate known AD variants for associations and underlying effects with other diseases as a starting point.

## Methods

### AD variants

A subset significantly associated to AD variants was identified using variants from the Alzheimer’s Disease Variant Portal (ADVP)^27^, a collection of variant associations harmonized from over 200 AD GWAS publications. Pulling literature from the Alzheimer’s Disease Genetics Consortium (ADGC) and GWAS Catalog, ADVP is an up-to-date resource of all AD genetic associations from studies of increasingly diverse populations and outcomes^27^. To identify reliably significant AD associated variants we subsetted a total of 1,863 variants from their original publications by a conservative p-value significance threshold of < 5*10^−8^. Ultimately 497 AD variants passed this threshold and were used going forward.

### Discovery in PheWAS Summary Statistics

Initial discovery was done using publicly available PheWAS summary statistics from an analysis using 361,194 genotypes (hg37) and 633 ICD10 code phenotypes from UK Biobank (UKBB) participants of European ancestry^28^. Summary statistics are located on PheWeb^28^. Variants were filtered based on minor allele frequency (MAF) > 0.1% and Hardy Weinburg equilibrium (HWE) > 1e-10, leaving 10,800,000 autosomal variants prior to analysis. Final summary statistics results were subsetted based on AD variants from ADVP, leaving 467 unique variants.

### Replication PheWAS in eMERGE Network

#### Genotypic Data

The eMERGE Network version III is made up of genotypic, lab and electronic health record information (ICD10 codes) from 105,108 individuals. The participants’ sex was 54.1% female (n=45268) and 45.9% male (n=38404) with an average age of 62.5 years. The group was made up of 75.9% European ancestry, 15.1% Black or African ancestry, 1.2% Asian ancestry, 0.2% American Indian or Alaskan Native, and 7.7% Unknown, Missing or Unreported ancestry. A subset of 83,672 participants containing demographic, genotype and phenotype information were used for our analysis. Genotype array data was provided by individual eMERGE center sites and imputed on genome build 37 (hg19) using HRC1.1 as an imputation reference^29^. Separate subject level and variant level missingness was filtered at a threshold of >2%. The eMERGE Phase-III data are available from dbGaP (dbGaP Study Accession: phs001584.v2.p2).

#### PheWAS replication

SAIGE^30^ was used to perform a GWAS for each phenotype in the eMERGE Network to control for case-control imbalance and sample relatedness. SAIGE uses a generalized mixed model to run associations between individual variants and phenotypes with case-control status^30^. ICD10 phenotypes were grouped by disease category with decimals removed. These ICD10 code groupings were then filtered for codes with >200 cases to ensure adequate power^31^. Ultimately 845 ICD10 code phenotypes were used for the PheWAS analysis. The step 1 model file for SAIGE was created using genotypes filtered with a MAF >0.05, HWE >0.05, and variant missingness <0.01.

### Variant impact on gene expression

Genes within 1MB of variants replicated in both cohorts were used to identify variant-gene pairs to consider as expression quantitative trait loci (eQTLs). Significance values for these eQTLs were then pulled in all available tissues in the Genotype-Tissue Expression (GTEx) portal (v8)^32^.

## Results

### Immune phenotypes are most frequent in AD variants

With 633 ICD10-based phenotypes and 467 AD variants in the UKBB discovery set, 81 significant associations passed the statistical significance threshold of p<5*10^−8^, primarily in cardiac and immune related diseases (Figure 2, Table 1). A PheWAS was also conducted in the eMERGE Network which included 845 ICD10-based phenotypes using the same AD variants. 71 variant-phenotype associations were found to be statistically significant independently in eMERGE (Figure 2, Table 2), with six of these associations also occurring in the UKBB discovery set (Table 3). The majority of significant associations in UKBB and eMERGE were phenotypes associated with variants located within the major histocompatibility complex region (chr6:28,477,797-33,448,354), which encodes clusters of genes involved in innate and acquired immune responses in humans^33^. The immune system is long believed to play a critical role in the progression of AD through prolonged activation and inflammation^34,35^.

**Table 1.** Top significant AD variants from UKBB.

| *rsID* | *MAF* | *P value* | *ICD10 code* | *Phenotype Description* | *AD GWAS (PMID)* |
| --- | --- | --- | --- | --- | --- |
| rs6931277 | 0.192298 | 3.375160e-98 | M06 | Other rheumatoid arthritis | 30617256 |
| rs198834 | 0.369300 | 9.760450e-85 | E83 | Disorders of mineral metabolism | 22685416 |
| rs115553053 | 0.000033 | 4.954240e-62 | L27 | Dermatitis due to substances taken internally | 23571587 |
| rs6931277 | 0.192298 | 1.417510e-52 | M05 | Rheumatoid arthritis with rheumatoid factor | 30617256 |
| rs9272561 | 0.340052 | 5.552970e-47 | K90 | Intestinal malabsorption | 30413934 |

**Table 2.** Significant variants from eMERGE.

| *rsID* | *MAF* | *P value* | *ICD10 code* | *Phenotype Description* | *AD GWAS (PMID)* |
| --- | --- | --- | --- | --- | --- |
| rs2516049 | 0.125072 | 6.981613e-37 | H52 | Disorders of refraction and accommodation | 27088644 |
| rs6931277 | 0.159755 | 4.700046e-36 | M05 | Rheumatoid arthritis with rheumatoid factor | 30617256 |
| rs6931277 | 0.159755 | 2.645999e-30 | E10 | Type 1 diabetes mellitus | 30617256 |
| rs6931277 | 0.159755 | 3.683423e-24 | M06 | Other rheumatoid arthritis | 30617256 |
| rs2516049 | 0.125072 | 3.459759e-22 | Z94 | Transplanted organ and tissue status | 27088644 |

**Table 3.** Variants showing significance in both UKBB and eMERGE.

| *rsID* | *Closest eQTL gene* | *ICD10 code* | *Phenotype Description* | *AD GWAS (PMID)* |
| --- | --- | --- | --- | --- |
| rs6931277 | *HLA-DQA1* | E10 | Type 1 diabetes mellitus | 30617256 |
| rs6931277 | *HLA-DQA1* | M06 | Other rheumatoid arthritis | 30617256 |
| rs9940128 | *FTO* | E66 | Overweight and obesity | 28870582 |
| rs6931277 | *HLA-DQA1* | M05 | Rheumatoid arthritis with rheumatoid factor | 30617256 |
| rs536810 | *HLA-DQA1* | M05 | Rheumatoid arthritis with rheumatoid factor | 30413934 |
| rs17747324 | *TCF7L2* | E11 | Type 2 diabetes mellitus | 30805717 |

**Figure 1.**
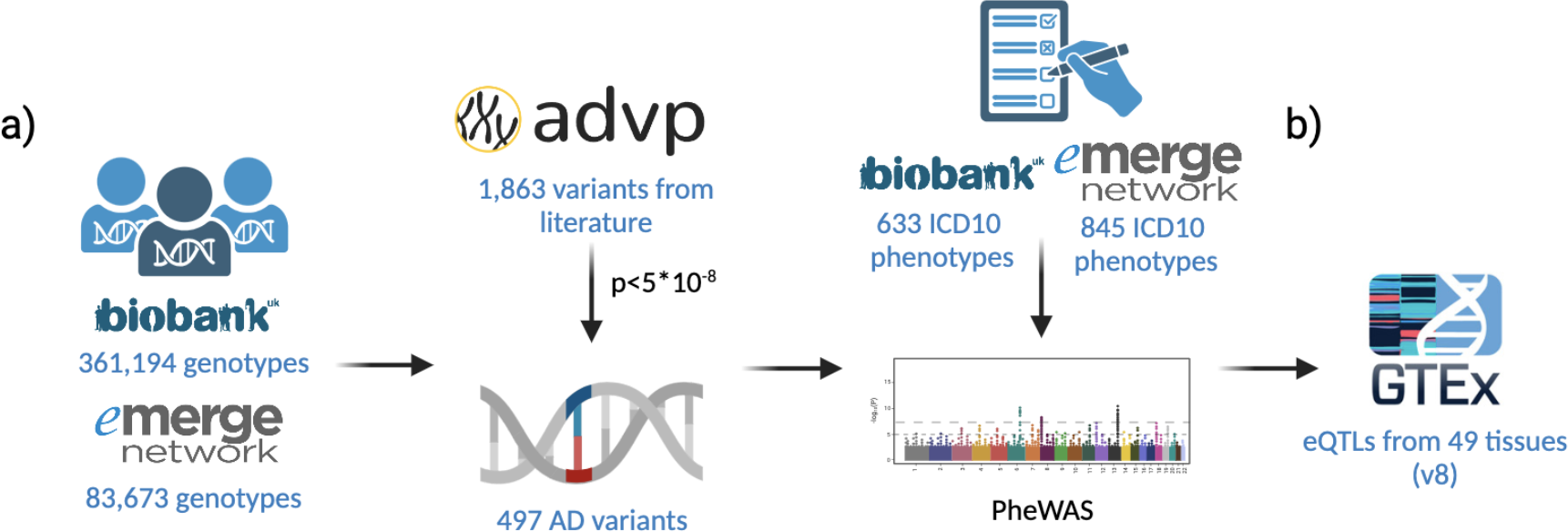
Overview of methods. a) Variants collected from the ADVP were subsetted to AD significant variants via p-value threshold of <5*10^−8^. These variants were subsetted from genotypes collected from the UKBB and eMERGE Network and used with ICD10-based phenotypes to conduct a PheWAS. b) Genes within 1MB of replicating significant variants were investigated using eQTLs in 49 tissues from GTEx (v8).

**Figure 2.**
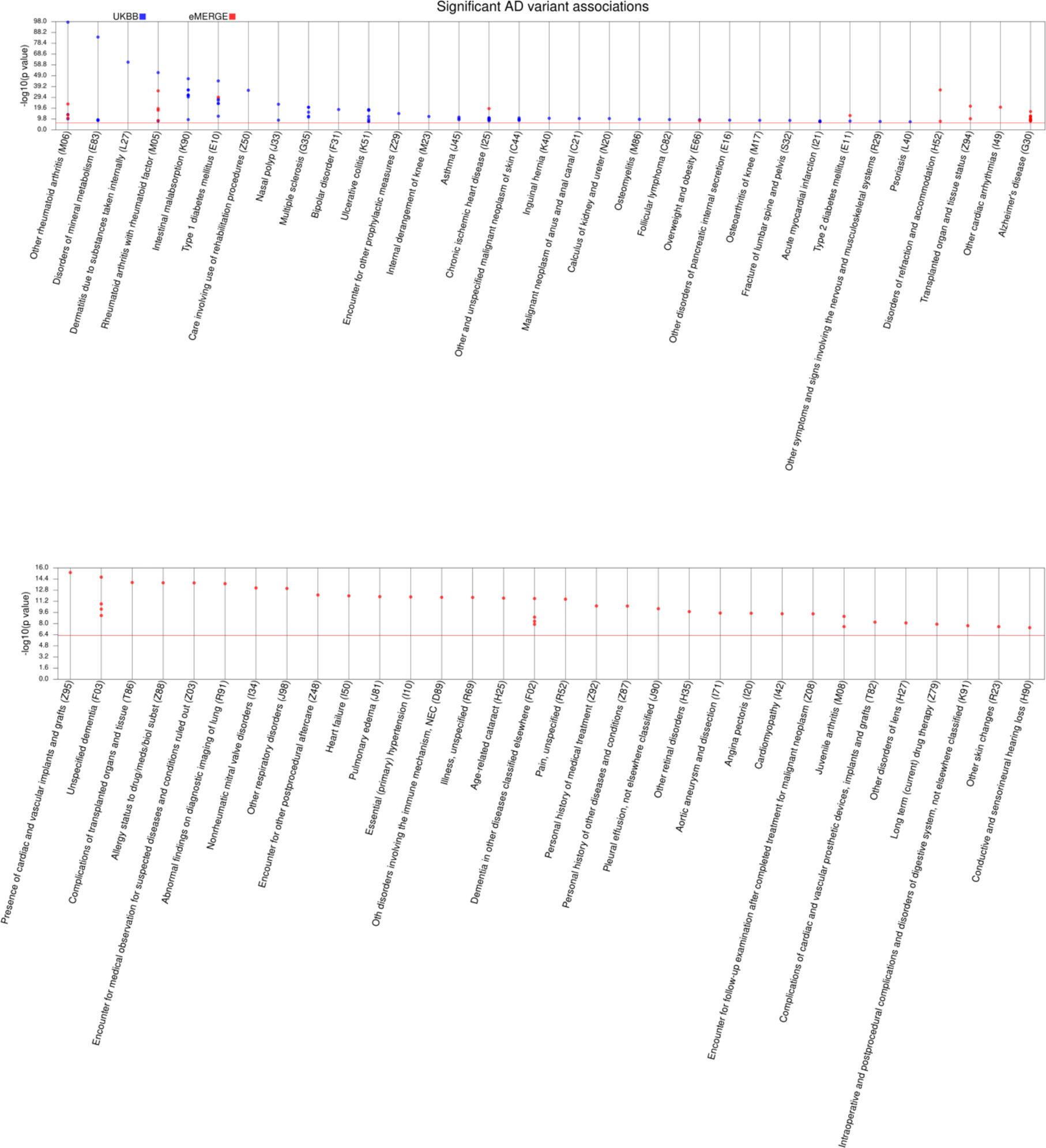
Significant results from both cohorts

#### MHC region

47 of the total 81 associations included variants that fall within the MHC region (Figure 3). Eight of these were significantly associated with intestinal malabsorption (K90) and seven variants were significantly associated with ulcerative colitis (K51) in the discovery set. Other diseases well believed to have overlap with AD appeared such as Type 1 diabetes (T1D) (E10), multiple sclerosis (MS) (G35) and rheumatoid arthritis (M05) also appeared to be significantly associated with at least one of the 467 AD variants (Supplementary Table 1). Type I diabetes was found to be associated with six different variants, some of which have been previously discussed for T1D such as rs6931277^36,37^. However rs9272561, rs9268877, rs9271058, rs9271192 and rs2516049 do not appear to previously show associations to T1D to the best of our knowledge. The theory that AD is primarily a metabolic disease is also supported by four variants located near or within the MHC region. Rs198834, rs2975033, rs9271192, rs9271058 all significantly associated with disorders of metabolic metabolism (E83).

**Figure 3.**
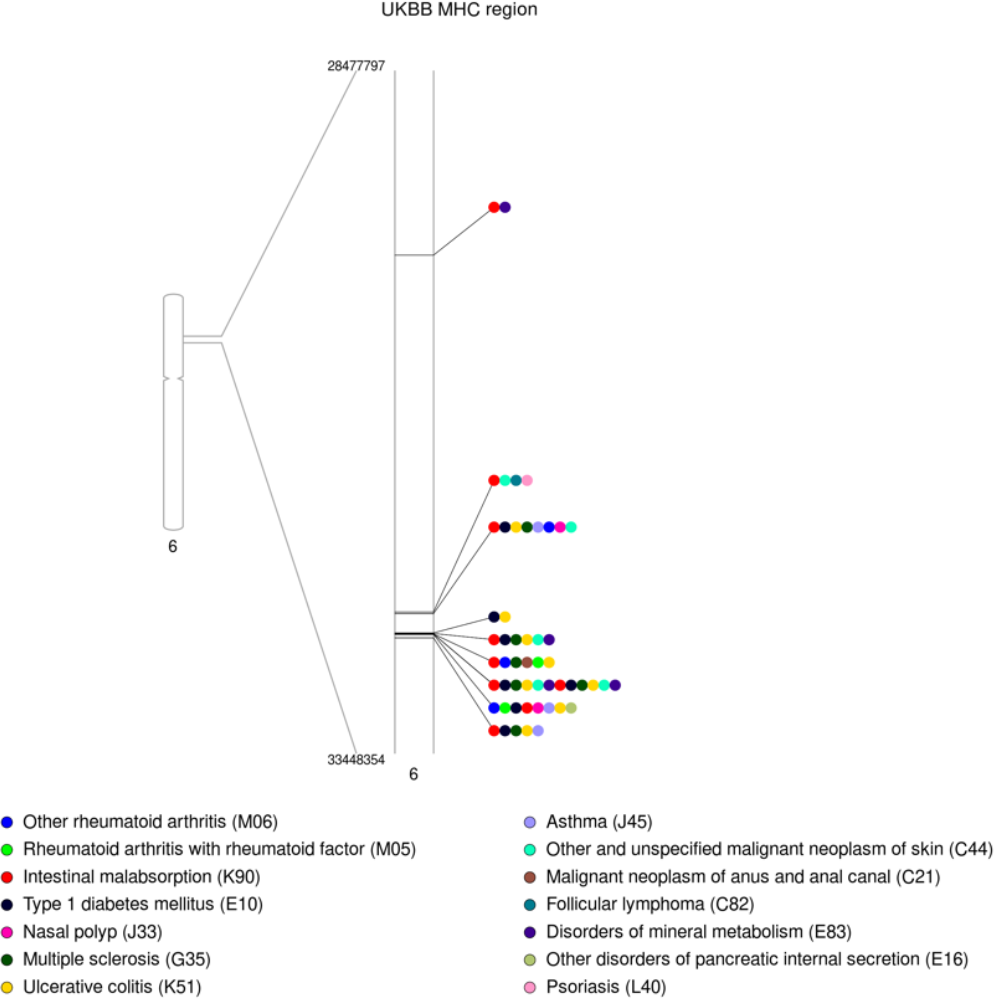
Significant variant associations within the MHC region from UKBB

#### Non-MHC region

34 associations included variants which fall outside of the MHC region, with most variants associating with chronic ischemic heart disease (14) (I25) and acute myocardial infarction (6) (I21) (Figure 4). Further, most of these associations were made up of variants that fall within 1MB up or downstream of the *APOE* gene, which is the most common gene linked to late-onset AD^38,39^. Two variants falling outside of the MHC region also replicated in eMERGE. Variant rs9940128 was significantly associated with overweight and obesity (E66) in both UKBB and eMERGE, as well as rs17747324 associating with T2D (E11).

**Figure 4.**
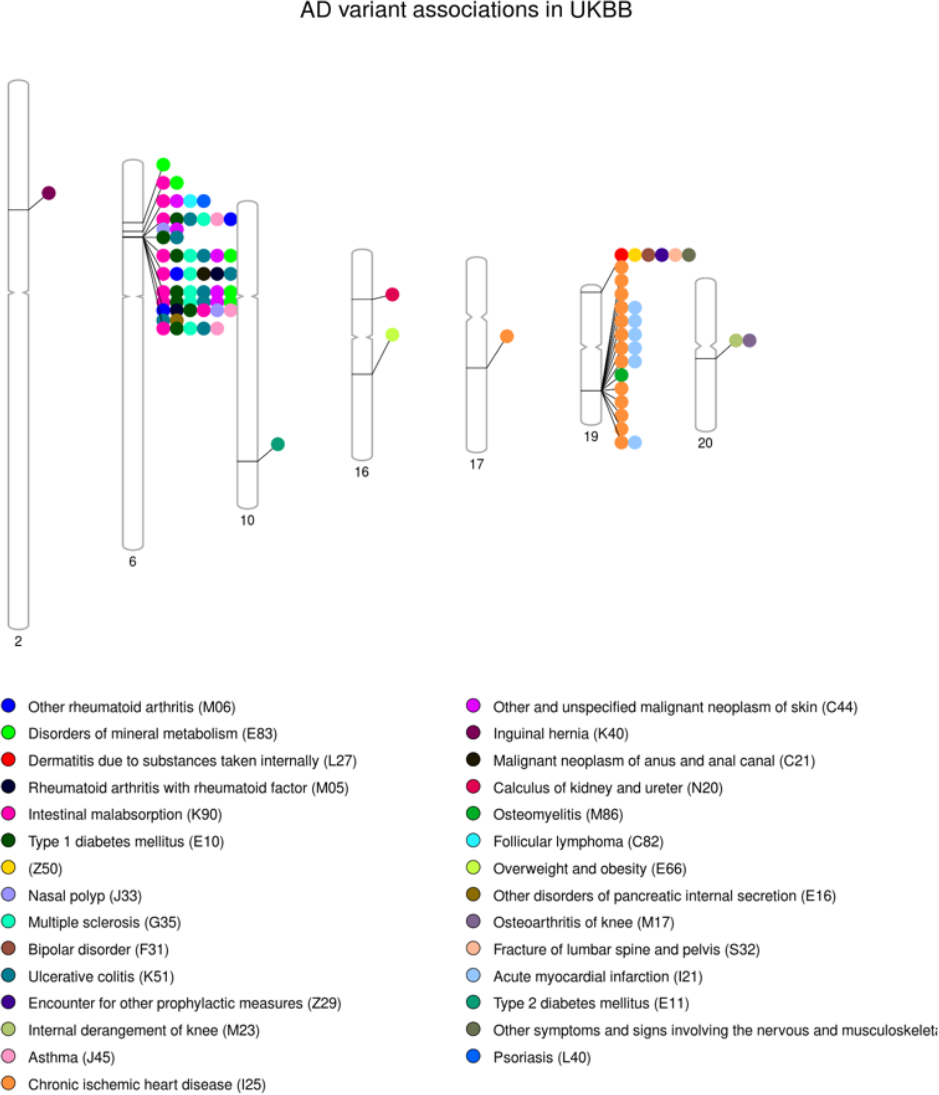
All significant variant associations from UKBB results

Four of these MHC-region associations replicated in the eMERGE PheWAS (Table 3), including rs6931277 which was significantly associated with T1D, rheumatoid arthritis with rheumatoid factor (M05), and other rheumatoid arthritis (M06) (Supplementary Table 2). Variant rs536810 also replicated with rheumatoid arthritis with rheumatoid factor in eMERGE.

### Variants impact gene expression in multiple tissues

Variants located within the MHC region tended to impact multiple *HLA* genes in multiple tissues (Supplementary Table 3). We pay special attention to eQTLs with significant values in brain tissues relevant to AD such as the frontal cortex, frontal cortex, and hippocampus^40,41^, as well as with tissues that may be relevant to the secondary associating phenotype. Of the replicating variants, rs6931277 and rs536810 had eQTLs with significant changes in gene expression in any brain tissue, including the frontal cortex and hippocampus. Both variants showed significant expression with *HLA-DQA2, HLA-DRB1*, and *HLA-DRB6* in brain tissues as well as relevant tissues to T1D and RA. For *HLA-DRB1* we see changes in the artery, heart, tibial nerve, pancreas, skin, and whole blood amongst others, in addition to changes in the majority of brain tissues with each variant. *HLA-DQA2* and *HLA-DRB6* showed widespread changes in expression with both rs6931277 and rs536810 in almost all tissues surveyed.

## Discussion

Most previous studies focused on AD genetics have been limited to disease specific cohorts of smaller numbers due to prevalence. However, our study aims to leverage previously confirmed AD variants, which allows us to bypass using AD-specific cohorts and take advantage of the entirety of large medical biobanks such as UKBB when looking for genetic associations with co-occurring disease. We used two biobanks, UKBB which included 361,194 genotypes of primarily European ancestry, along with 633 ICD10-based phenotypes, far larger than any AD cohort study to date. We replicated this with a second large, more diverse cohort from the eMERGE Network, made up of 83, 672 genotypes tested against 845 ICD10-based phenotypes.

We observed 81 unique variant-phenotype associations within UKBB (Table 1), 71 associations detected within eMERGE (Table 2), and six associations that overlapped in both cohorts (Table 3). Most (47 UKBB and 45 eMERGE) significant variant associations from both cohorts include variants located within the MHC or human leukocyte antigen (HLA) region which is a complex on chromosome 6 made up of genes responsible for regulation of the immune system. The immune system is known to play a major role in AD pathology within the brain. Activated microglia and astrocytes respond to amyloid beta (Aβ) protein build up by releasing pro-inflammatory cytokines which stimulates a chronic inflammatory reaction^42^. These inflammatory molecules can result in neuronal cell death as well as promoting wider effects in the body by spreading via a weakened blood brain barrier (BBB)^43^.

Within UKBB, eight individual AD variants (rs9272561, p=5.55e^-47^; rs9268877, p=6.71e^-37^; rs115674098, p=8.39e^-37^; rs6931277, p=2.50e^-32^; rs9271192, p=3.98e^-32^; rs9271058, p=7.07e^-32^; rs536810, p=1.58e^-30^; rs2975033, p=5.00e^-10^) were associated with intestinal malabsorption and seven variants (rs9268877, p=3.86e^-19^; rs9271058, p=1.79e^-18^; rs9271192, p=1.90e^-18^; rs9272561, p=7.69e^-13^; rs6931277, p=2.58e^-10^; rs536810, p=1.65e^-08^; rs2516049, p=2.42e^-08^) significantly associated with ulcerative colitis, both of which can be grouped as gut disorders. The connection between the gut microbiome, inflammation and AD has become increasingly of interest, with more focus being given to the implications of the gut-brain axis. This bidirectional communication between gut microbiota and brain occurs via the immune and nervous system to maintain bodily homeostasis^44^. Changes in microbiota composition are now believed to contribute to AD pathogenesis^45,46^ and diseases like Crohn’s disease and inflammatory bowel disease have previously been associated with greater AD risk^47,48^.

Top eMERGE results within the MHC region were dominated by associations with rheumatoid arthritis (RA) with rheumatoid factor (rs6931277, p=4.70e^-36^; rs115124923, p=6.30e^-20^; rs2516049, p=1.66e^-18^; rs536810, p=3.43e^-09^) other RA (rs6931277, p=3.68e^-24^; rs2516049, p=2.49e^-14^; rs115124923, p=2.36e^-11^), and juvenile arthritis (rs2516049, p=9.30e^-10^; rs115124923, p=2.73e^-08^). To our knowledge rs115124923 has yet to be associated with any form of arthritis, and yet it is significantly associated with three forms in eMERGE. RA is also traced back to the immune system and chronic inflammation, where inflammatory cells are activated and recruited to joints where they incite damage^49^. Patients with RA have shown to have an increased risk for AD, whereby treatment to reduce inflammation for RA via anti-TNF agents decreased risk of AD^50^. Two variants replicated in both cohorts for RA, rs6931277 for RA with rheumatoid factor and other RA along with rs536810 and RA with rheumatoid factor.

Outside of the MHC region we also saw a high instance of cardiac-related phenotypes. The most frequent associating phenotypes outside of the MHC region in UKBB were chronic ischemic heart disease (14 variants) and acute myocardial infarction (6 variants), with almost all of these tracing back to within 1MB of the well-known AD risk gene *APOE* (chr19:44,905,791-44,909,393). *APOE* modulates lipoprotein and Aβ metabolism and has been well documented as a risk factor for both AD and cardiac-related diseases^51,52^. We did not see replication of these associations in eMERGE.

To consider effects of variants at the gene level, we collected eQTLs for genes nearby significant variants to identify tissues in which they may be relevant, especially for eQTLs with significant p values in brain tissues pertinent to AD pathology (cortex, hippocampus), as well as relevant secondary disease tissues. However, to formally test for pleiotropic effects within tissues relevant to AD and the secondary disease, we plan to finemap and colocalize significant PheWAS signals directly with eQTLs in tissues from GTEx. It is also worth noting that our study was limited by phenotyping considerations as well as sample sizes for certain phenotypes. By using disease categories instead of full disease codes we may be limiting the specificity of our associations. Additionally, given our threshold of 200 cases per phenotype to maintain power, several diagnosis codes were left out, and not all diagnosis codes overlapped between the two tested cohorts. As part of future work we will further replicate these results in the Penn Medicine BioBank (PMBB) to apply an additional diverse population set.

## Conclusion

In summary, we provide a starting point for investigating broad impacts that variants associated with AD may have on disease progression. In addition to variants already known to be associated with other diseases, like rs6931277 and T1D^36,37^, we add AD variants with new secondary associations. UKBB showed variant associations with intestinal disorder phenotypes, and eMERGE showed new variant associations with RA. While we plan to further validate and expand these findings in additional datasets, this study demonstrates the potential of using PheWAS methods to identify promising target areas for drug repurposing, as well as better understanding key contributors to overall AD progression.

## Data Availability

All data used in this manuscript are available online: UKBB (https://www.ukbiobank.ac.uk/enable-your-research/apply-for-access), eMERGEIII (dbGaP Study Accession: phs001584.v2.p2)
All data produced in the present study are available upon reasonable request to the authors.

## Acknowledgements

This work was supported in part by R01HG010067, R01GM138597, R01AG066833.

We would like to acknowledge the following members of the eMERGE Network: Yogasudha Veturi, PhD (Pennsylvania State University), Anastasia Lucas (University of Pennsylvania), Yuki Bradford, MS (University of Pennsylvania), Anurag Verma, PhD (University of Pennsylvania), Shefali Setia Verma, PhD (University of Pennsylvania), Joseph Park, PhD (University of Pennsylvania), Wei-Qi Wei, MD, PhD (Vanderbilt University Medical Center), Bahram Namjou-Khales, MD (Cincinnati Children’s Hospital), Krzysztof Kiryluk, MD, MS (Columbia University), Iftikhar Kullo, MD (Mayo Foundation), Yuan Luo, PhD (Northwestern University), QiPing Feng, PhD (Vanderbilt University Medical Center), Binglan Li, PhD (Stanford University).

The eMERGE Network was initiated and funded by NHGRI through the following grants:

Phase III: U01HG8657 (Kaiser Permanente Washington (formerly known at GroupHealth) /University of Washington); U01HG8685 (Brigham and Women’s Hospital); U01HG8672 (Vanderbilt University Medical Center); U01HG8666 (Cincinnati Children’s Hospital Medical Center); U01HG6379 (Mayo Clinic); U01HG8679 (Geisinger Clinic); U01HG8680 (Columbia University Health Sciences); U01HG8684 (Children’s Hospital of Philadelphia); U01HG8673 (Northwestern University); U01HG8701 (Vanderbilt University Medical Center serving as the Coordinating Center); U01HG8676 (Partners Healthcare/Broad Institute); and U01HG8664 (Baylor College of Medicine)

Phase II: U01HG006828 (Cincinnati Children’s Hospital Medical Center/Boston Children’s Hospital); U01HG006830 (Children’s Hospital of Philadelphia); U01HG006389 (Essentia Institute of Rural Health, Marshfield Clinic Research Foundation and Pennsylvania State University); U01HG006382 (Geisinger Clinic); U01HG006375 (Group Health Cooperative/University of Washington); U01HG006379 (Mayo Clinic); U01HG006380 (Icahn School of Medicine at Mount Sinai); U01HG006388 (Northwestern University); U01HG006378 (Vanderbilt University Medical Center); and U01HG006385 (Vanderbilt University Medical Center serving as the Coordinating Center).

If the project includes data from the eMERGE imputed merged Phase I and Phase II dataset, please also add U01HG004438 (CIDR) and U01HG004424 (the Broad Institute) serving as Genotyping Centers. And/or The PGRNSeq dataset (eMERGE PGx), please also add U01HG004438 (CIDR) serving as a Sequencing Center.

Phase I: U01-HG-004610 (Kaiser Permanente Washington /University of Washington); U01-HG-004608 (Marshfield Clinic Research Foundation and Vanderbilt University Medical Center); U01-HG-04599 (Mayo Clinic); U01HG004609 (Northwestern University); U01-HG-04603 (Vanderbilt University Medical Center, also serving as the Administrative Coordinating Center); U01HG004438 (CIDR) and U01HG004424 (the Broad Institute) serving as Genotyping Centers.

## References

1. 2023 Alzheimer’s disease facts and figures. Alzheimers Dement [Internet]. 2023 Mar 14; Available from: 10.1002/alz.13016

2. Glenner GG, Wong CW. Alzheimer’s disease: initial report of the purification and characterization of a novel cerebrovascular amyloid protein. Biochem Biophys Res Commun. 1984 May 16;120(3):885–90.

3. Ittner LM, Ke YD, Delerue F, Bi M, Gladbach A, van Eersel J, et al. Dendritic function of tau mediates amyloid-beta toxicity in Alzheimer’s disease mouse models. Cell. 2010 Aug 6;142(3):387–97.

4. Kar S, Slowikowski SPM, Westaway D, Mount HTJ. Interactions between beta-amyloid and central cholinergic neurons: implications for Alzheimer’s disease. J Psychiatry Neurosci. 2004 Nov;29(6):427–41.

5. Gatz M, Reynolds CA, Fratiglioni L, Johansson B, Mortimer JA, Berg S, et al. Role of genes and environments for explaining Alzheimer disease. Arch Gen Psychiatry. 2006 Feb;63(2):168–74.

6. Lloret A, Esteve D, Lloret MA, Cervera-Ferri A, Lopez B, Nepomuceno M, et al. When Does Alzheimer’s Disease Really Start? The Role of Biomarkers. Focus. 2021 Jul;19(3):355–64.

7. McDade E, Bateman RJ. Stop Alzheimer’s before it starts. Nature. 2017 Jul 12;547(7662):153–5.

8. Tanzi RE. The genetics of Alzheimer disease. Cold Spring Harb Perspect Med [Internet]. 2012 Oct 1;2(10). Available from: 10.1101/cshperspect.a006296

9. Tesi N, van der Lee SJ, Hulsman M, Jansen IE, Stringa N, van Schoor NM, et al. Immune response and endocytosis pathways are associated with the resilience against Alzheimer’s disease. Transl Psychiatry. 2020 Sep 29;10(1):332.

10. de la Monte SM, Tong M. Brain metabolic dysfunction at the core of Alzheimer’s disease. Biochem Pharmacol. 2014 Apr 15;88(4):548–59.

11. Kao YC, Ho PC, Tu YK, Jou IM, Tsai KJ. Lipids and Alzheimer’s Disease. Int J Mol Sci [Internet]. 2020 Feb 22;21(4). Available from: 10.3390/ijms21041505

12. Wang XF, Lin X, Li DY, Zhou R, Greenbaum J, Chen YC, et al. Linking Alzheimer’s disease and type 2 diabetes: Novel shared susceptibility genes detected by cFDR approach. J Neurol Sci. 2017 Sep 15;380:262–72.

13. Lowell BB, Shulman GI. Mitochondrial dysfunction and type 2 diabetes. Science. 2005 Jan 21;307(5708):384–7.

14. De Felice FG, Lourenco MV, Ferreira ST. How does brain insulin resistance develop in Alzheimer’s disease? Alzheimers Dement. 2014 Feb;10(1 Suppl):S26–32.

15. Newman AB, Fitzpatrick AL, Lopez O, Jackson S, Lyketsos C, Jagust W, et al. Dementia and Alzheimer’s disease incidence in relationship to cardiovascular disease in the Cardiovascular Health Study cohort. J Am Geriatr Soc. 2005 Jul;53(7):1101–7.

16. Rosenberg PB, Mielke MM, Tschanz J, Cook L, Corcoran C, Hayden KM, et al. Effects of cardiovascular medications on rate of functional decline in Alzheimer disease. Am J Geriatr Psychiatry. 2008 Nov;16(11):883–92.

17. Khachaturian AS, Zandi PP, Lyketsos CG, Hayden KM, Skoog I, Norton MC, et al. Antihypertensive medication use and incident Alzheimer disease: the Cache County Study. Arch Neurol. 2006 May;63(5):686–92.

18. Janson J, Laedtke T, Parisi JE, O’Brien P, Petersen RC, Butler PC. Increased risk of type 2 diabetes in Alzheimer disease. Diabetes. 2004 Feb;53(2):474–81.

19. Wang C, Holtzman DM. Bidirectional relationship between sleep and Alzheimer’s disease: role of amyloid, tau, and other factors. Neuropsychopharmacology. 2020 Jan;45(1):104–20.

20. Ju YES, McLeland JS, Toedebusch CD, Xiong C, Fagan AM, Duntley SP, et al. Sleep quality and preclinical Alzheimer disease. JAMA Neurol. 2013 May;70(5):587–93.

21. Guarnieri B, Adorni F, Musicco M, Appollonio I, Bonanni E, Caffarra P, et al. Prevalence of sleep disturbances in mild cognitive impairment and dementing disorders: a multicenter Italian clinical cross-sectional study on 431 patients. Dement Geriatr Cogn Disord. 2012 Mar 8;33(1):50–8.

22. Bone WP, Siewert KM, Jha A, Klarin D, Damrauer SM, VA Million Veteran Program, et al. Multi-trait association studies discover pleiotropic loci between Alzheimer’s disease and cardiometabolic traits. Alzheimers Res Ther. 2021 Feb 4;13(1):34.

23. Paik H, Lee J, Jeong CS, Park JS, Lee JH, Rappoport N, et al. Identification of a pleiotropic effect of ADIPOQ on cardiac dysfunction and Alzheimer’s disease based on genetic evidence and health care records. Transl Psychiatry. 2022 Sep 16;12(1):389.

24. Craft S, Claxton A, Baker LD, Hanson AJ, Cholerton B, Trittschuh EH, et al. Effects of Regular and Long-Acting Insulin on Cognition and Alzheimer’s Disease Biomarkers: A Pilot Clinical Trial. J Alzheimers Dis. 2017;57(4):1325–34.

25. Craft S, Baker LD, Montine TJ, Minoshima S, Watson GS, Claxton A, et al. Intranasal insulin therapy for Alzheimer disease and amnestic mild cognitive impairment: a pilot clinical trial. Arch Neurol. 2012 Jan;69(1):29–38.

26. Claxton A, Baker LD, Hanson A, Trittschuh EH, Cholerton B, Morgan A, et al. Long-acting intranasal insulin detemir improves cognition for adults with mild cognitive impairment or early-stage Alzheimer’s disease dementia. J Alzheimers Dis. 2015;44(3):897–906.

27. Kuksa PP, Liu CL, Fu W, Qu L, Zhao Y, Katanic Z, et al. Alzheimer’s Disease Variant Portal: A Catalog of Genetic Findings for Alzheimer’s Disease. J Alzheimers Dis. 2022;86(1):461–77.

28. Howrigan DP, Abbott L, rkwalters, Palmer D, Francioli L, Hammerbacher J. Nealelab/UK_Biobank_GWAS: v2 [Internet]. Zenodo; 2023. Available from: https://zenodo.org/record/8011557

29. Stanaway IB, Hall TO, Rosenthal EA, Palmer M, Naranbhai V, Knevel R, et al. The eMERGE genotype set of 83,717 subjects imputed to ∼40 million variants genome wide and association with the herpes zoster medical record phenotype. Genet Epidemiol. 2019 Feb;43(1):63–81.

30. Zhou W, Nielsen JB, Fritsche LG, Dey R, Gabrielsen ME, Wolford BN, et al. Efficiently controlling for case-control imbalance and sample relatedness in large-scale genetic association studies. Nat Genet. 2018 Sep;50(9):1335–41.

31. Verma A, Bradford Y, Dudek S, Lucas AM, Verma SS, Pendergrass SA, et al. A simulation study investigating power estimates in phenome-wide association studies. BMC Bioinformatics. 2018 Apr 4;19(1):120.

32. GTEx Consortium. The GTEx Consortium atlas of genetic regulatory effects across human tissues. Science. 2020 Sep 11;369(6509):1318–30.

33. Horton R, Wilming L, Rand V, Lovering RC, Bruford EA, Khodiyar VK, et al. Gene map of the extended human MHC. Nat Rev Genet. 2004 Dec;5(12):889–99.

34. Zhang R, Miller RG, Madison C, Jin X, Honrada R, Harris W, et al. Systemic immune system alterations in early stages of Alzheimer’s disease. J Neuroimmunol. 2013 Mar 15;256(1-2):38–42.

35. McGeer PL, Akiyama H, Itagaki S, McGeer EG. Immune system response in Alzheimer’s disease. Can J Neurol Sci. 1989 Nov;16(4 Suppl):516–27.

36. Nguyen C, Varney MD, Harrison LC, Morahan G. Definition of high-risk type 1 diabetes HLA-DR and HLA-DQ types using only three single nucleotide polymorphisms. Diabetes. 2013 Jun;62(6):2135–40.

37. Brorsson CA, Onengut S, Chen WM, Wenzlau J, Yu L, Baker P, et al. Novel Association Between Immune-Mediated Susceptibility Loci and Persistent Autoantibody Positivity in Type 1 Diabetes. Diabetes. 2015 Aug;64(8):3017–27.

38. Liu CC, Liu CC, Kanekiyo T, Xu H, Bu G. Apolipoprotein E and Alzheimer disease: risk, mechanisms and therapy. Nat Rev Neurol. 2013 Feb;9(2):106–18.

39. Meyer MR, Tschanz JT, Norton MC, Welsh-Bohmer KA, Steffens DC, Wyse BW, et al. APOE genotype predicts when--not whether--one is predisposed to develop Alzheimer disease. Nat Genet. 1998 Aug;19(4):321–2.

40. Wenk GL. Neuropathologic changes in Alzheimer’s disease. J Clin Psychiatry. 2003;64 Suppl 9:7–10.

41. Bäckman L, Andersson JL, Nyberg L, Winblad B, Nordberg A, Almkvist O. Brain regions associated with episodic retrieval in normal aging and Alzheimer’s disease. Neurology. 1999 Jun 10;52(9):1861–70.

42. Serpente M, Bonsi R, Scarpini E, Galimberti D. Innate immune system and inflammation in Alzheimer’s disease: from pathogenesis to treatment. Neuroimmunomodulation. 2014 Feb 14;21(2-3):79–87.

43. Newcombe EA, Camats-Perna J, Silva ML, Valmas N, Huat TJ, Medeiros R. Inflammation: the link between comorbidities, genetics, and Alzheimer’s disease. J Neuroinflammation. 2018 Sep 24;15(1):276.

44. Cryan JF, O’Riordan KJ, Cowan CSM, Sandhu KV, Bastiaanssen TFS, Boehme M, et al. The Microbiota-Gut-Brain Axis. Physiol Rev. 2019 Oct 1;99(4):1877–2013.

45. Sochocka M, Donskow-Łysoniewska K, Diniz BS, Kurpas D, Brzozowska E, Leszek J. The Gut Microbiome Alterations and Inflammation-Driven Pathogenesis of Alzheimer’s Disease-a Critical Review. Mol Neurobiol. 2019 Mar;56(3):1841–51.

46. Vogt NM, Kerby RL, Dill-McFarland KA, Harding SJ, Merluzzi AP, Johnson SC, et al. Gut microbiome alterations in Alzheimer’s disease. Sci Rep. 2017 Oct 19;7(1):13537.

47. Szandruk-Bender M, Wiatrak B, Szeląg A. The Risk of Developing Alzheimer’s Disease and Parkinson’s Disease in Patients with Inflammatory Bowel Disease: A Meta-Analysis. J Clin Med Res [Internet]. 2022 Jun 27;11(13). Available from: 10.3390/jcm11133704

48. Aggarwal M, Alkhayyat M, Abou Saleh M, Sarmini MT, Singh A, Garg R, et al. Alzheimer Disease Occurs More Frequently In Patients With Inflammatory Bowel Disease: Insight From a Nationwide Study. J Clin Gastroenterol. 2023;57(5):501–7.

49. Firestein GS. Evolving concepts of rheumatoid arthritis. Nature. 2003 May 15;423(6937):356–61.

50. Chou RC, Kane M, Ghimire S, Gautam S, Gui J. Treatment for Rheumatoid Arthritis and Risk of Alzheimer’s Disease: A Nested Case-Control Analysis. CNS Drugs. 2016 Nov;30(11):1111–20.

51. Eichner JE, Dunn ST, Perveen G, Thompson DM, Stewart KE, Stroehla BC. Apolipoprotein E polymorphism and cardiovascular disease: a HuGE review. Am J Epidemiol. 2002 Mar 15;155(6):487–95.

52. Tiret L, de Knijff P, Menzel HJ, Ehnholm C, Nicaud V, Havekes LM. ApoE polymorphism and predisposition to coronary heart disease in youths of different European populations. The EARS Study. European Atherosclerosis Research Study. Arterioscler Thromb. 1994 Oct;14(10):1617–24.

